# NTT Docomo and Apple mobility data compared as countermeasures against COVID-19 outbreak in Japan

**DOI:** 10.1101/2020.05.01.20087155

**Authors:** Yoshiyuki Sugishita, Junko Kurita, Tamie Sugawara, Yasushi Ohkusa

## Abstract

**Background:** In Japan, as a measure to inhibit the COVID-19 outbreak, voluntary restrictions against going out (VRG) have been applied.

**Object:** Mobility information provided by Apple Inc. and NTT Docomo were assessed in terms of its usefulness in predicting conditions exacerbating an outbreak.

**Method:** A polynomial function was applied to daily Apple and Docomo data to calculate the observed R(t).

**Results:** The correlation coefficient among Apple and Docomo data was 0.91. The adjusted coefficient of determination for R(t) for the whole study period was higher using Docomo data than when Apple data were used. When we regressed R(t) on daily Apple and Docomo data simultaneously, the estimated coefficient of Docomo data was not significant.

**Discussion and Conclusion:** We demonstrated that Apple mobility data might be superior to Docomo data for explaining the entire course of the COVID-19 outbreak in Japan.

## Introduction

To support planning and evaluation, some methods to avoid and overcome COVID-19 outbreak peaks can be inferred. Those two methods are herd immunity [1] and infection countermeasures [2].

In Japan, in preference to lockdowns such as those mandated in European and North American countries, voluntary restrictions against going out (VRG) were announced by national and local governments from the end of March [3]. However, the VRG program intensity has changed over time. Public cooperation with VRG has also been changing. Lockdowns such as those instituted by the respective governments in Europe and North America might eventually engender cessation of widespread infection. However, in Japan, efforts are voluntary: people can adjust their own degree of cooperation independently. Therefore, the government should monitor current outbreak phenomena when moderating requirements for VRG. One difficulty of such monitoring efforts is that reporting the number of the newly infected patients entails some delay. One delay is attributable to the incubation period from infection to onset; another delay is that occurring after onset until reporting. Taken together, for approximately two weeks, one is unable to observe a precise daily number of newly infected persons. Therefore, because the latest information might not be timely, policies might be less effective if a government waits two weeks to conduct decision-making.

Data for many variables monitored for decision making have been reported from several services, including those of Apple Inc. and Alphabet Inc. (hereinafter Apple and Google, respectively) worldwide, and Nippon Telegraph and Telephone (NTT) Docomo (hereinafter Docomo) and East and West Japan Railway companies (JR) in Japan. Of those companies, Apple and Docomo started to publish related daily data from January [6,7]. Docomo data were used as an index of VRG by the government. Then one question must be raised: Which dataset can explain the outbreak better? The present study compares two data sources from the perspective of their power to predict outbreaks.

Going out was defined as a route search on Apple map using Apple data. Therefore, it excludes going out without searching for a well known place; it also incorrectly includes searches done at home. By contrast, Docomo data are based on the mobile phone location, as measured by 500 m grid. Therefore, it might be a more appropriate definition of going out.

## Methods

We first estimate the effective reproduction number R(t) n Japan assuming an incubation period following the empirical distribution in Japan. The number of symptomatic patients reported by the Ministry of Labour, Health and Welfare (MLHW) for January 14 – May 25 published [7] on May 27 were used. Some patients were excluded from data: those presumed to be persons infected abroad or infected as passengers on the Diamond Princess. Those patients were presumed not to represent community-acquired infection in Japan.

For the onset dates of some symptomatic patients that were unknown, we estimated their onset date from an empirical distribution with durations extending from onset to the report date among patients for whom the onset date had been reported. We estimated the onset date of patients for whom onset dates were not reported as follows: Letting *f*(*k*) represent this empirical distribution and letting *N*_*t*_ denote the number of patients for whom onset dates were not published by date *t*, then the number of patients for whom the onset date was known is *t*-1. The number of patients for whom onset dates were not available was estimated as *f*(1)*N*_*t*_. Similarly, the number of patients with onset date *t*-2 and for whom onset dates were not available was estimated as *f*(2)*N*_*t*_. Therefore, the total number of patients for whom the onset date was not available, given an onset date of *s*, was estimated as Σ_*k*_=1*f*(*k*)*N*_*s*_+*k* for the long duration extending from *s*.

Moreover, the reporting delay for published data from MHLW might be considerable. In other words, if *s*+*k* is larger than that in the current period *t*, then *s*+*k* represents the future for period *t*. For that reason, *Ns*+*k* is not observable. Such a reporting delay engenders underestimation bias of the number of patients. For that reason, it must be adjusted as 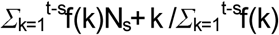 Similarly, patients for whom the onset dates were available are expected to be affected by the reporting delay. Therefore, we have 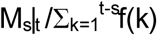, where *M*_*s*_|*t* represents the reported number of patients for whom onset dates were within period *s*, extending until the current period *t*.

We defined R(*t*) as the number of infected patients on day *t* divided by the number of patients who were presumed to be infectious. The number of infected patients was calculated from the epidemic curve by the onset date using a distribution of the incubation period. The distribution of infectiousness in symptomatic and asymptomatic cases was assumed to be 30% on the onset day, 20% on the following day, and 10% for the subsequent five days [8].

To clarify associations among the R(t) and mobility data, we regressed R(t) on a polynomial function of daily Apple and Docomo data, respectively. Moreover, we regressed R(t) simultaneously on a polynomial function of daily Apple and Docomo data. In any case, the order of the polynomial function was selected stepwise while all coefficients were significant.

The study period was February 10 through May 25. Moreover, we analyzed only those data recorded after March 10. In both cases, the data used for regression were those up to the end of April. Then we evaluated the predictive power in May.

## Ethical consideration

All information used for this study has been published [6–8]. Therefore there is no ethical issue related to this study.

## Results

As of May 27, using data for January 14 – May 25 in Japan, 14,972 community-acquired cases were identified, excluding asymptomatic cases. Figure 1 presents an empirical distribution of the duration of onset to reporting in Japan. The maximum delay was 30 days. Figure 2 depicts the empirical distribution of incubation periods among 125 cases for which the exposed date and onset date were published by MHLW in Japan. The mode was six days. The average was 6.6 days.

**Figure 1:**
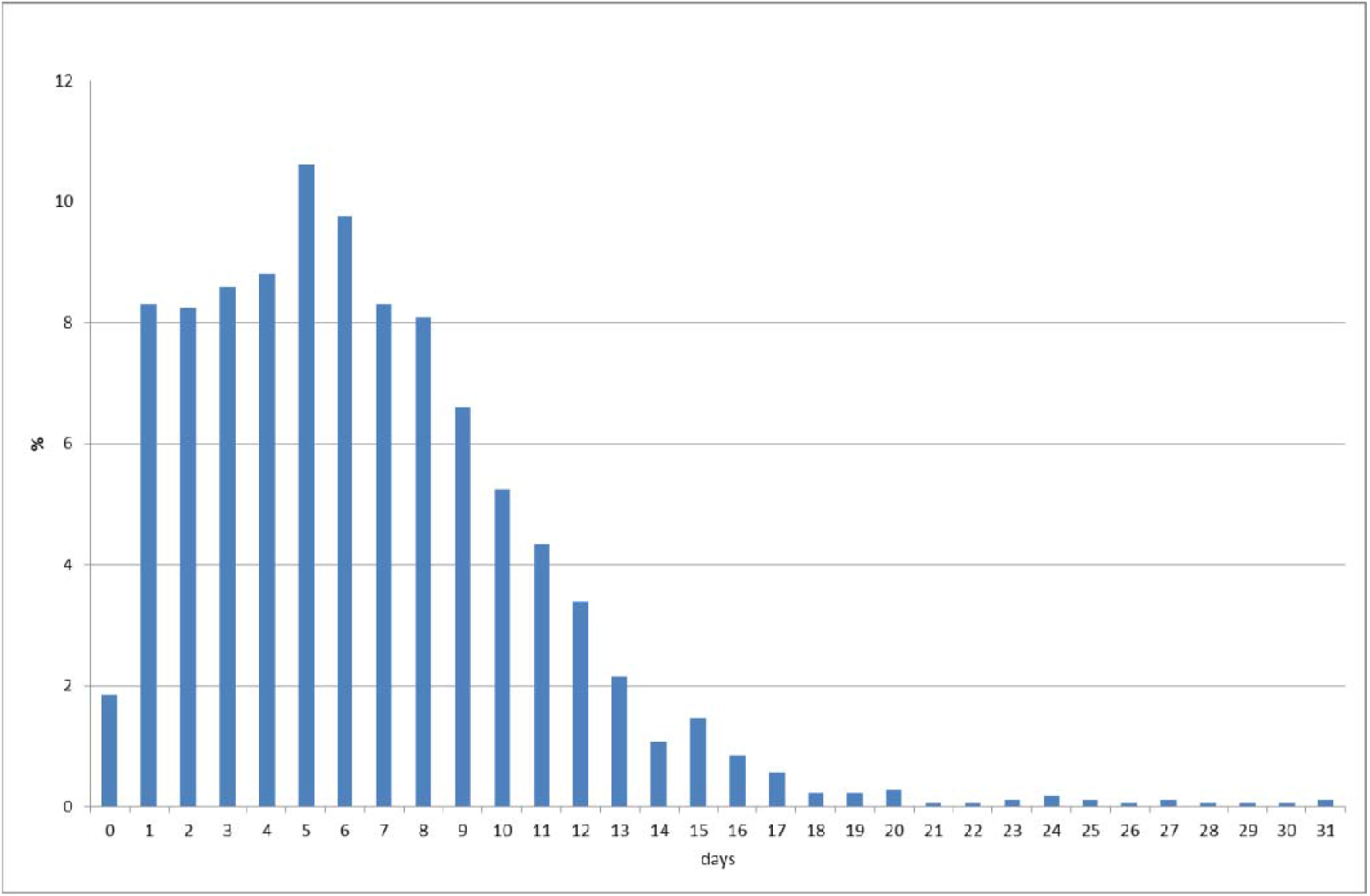
Empirical distribution of duration from onset to report by MLHW, Japan. Note: Bars represent the probability of duration from onset to report based on 657 patients for whom the onset date was available in Japan. Data were obtained from MLHW, Japan.

**Figure 2:**
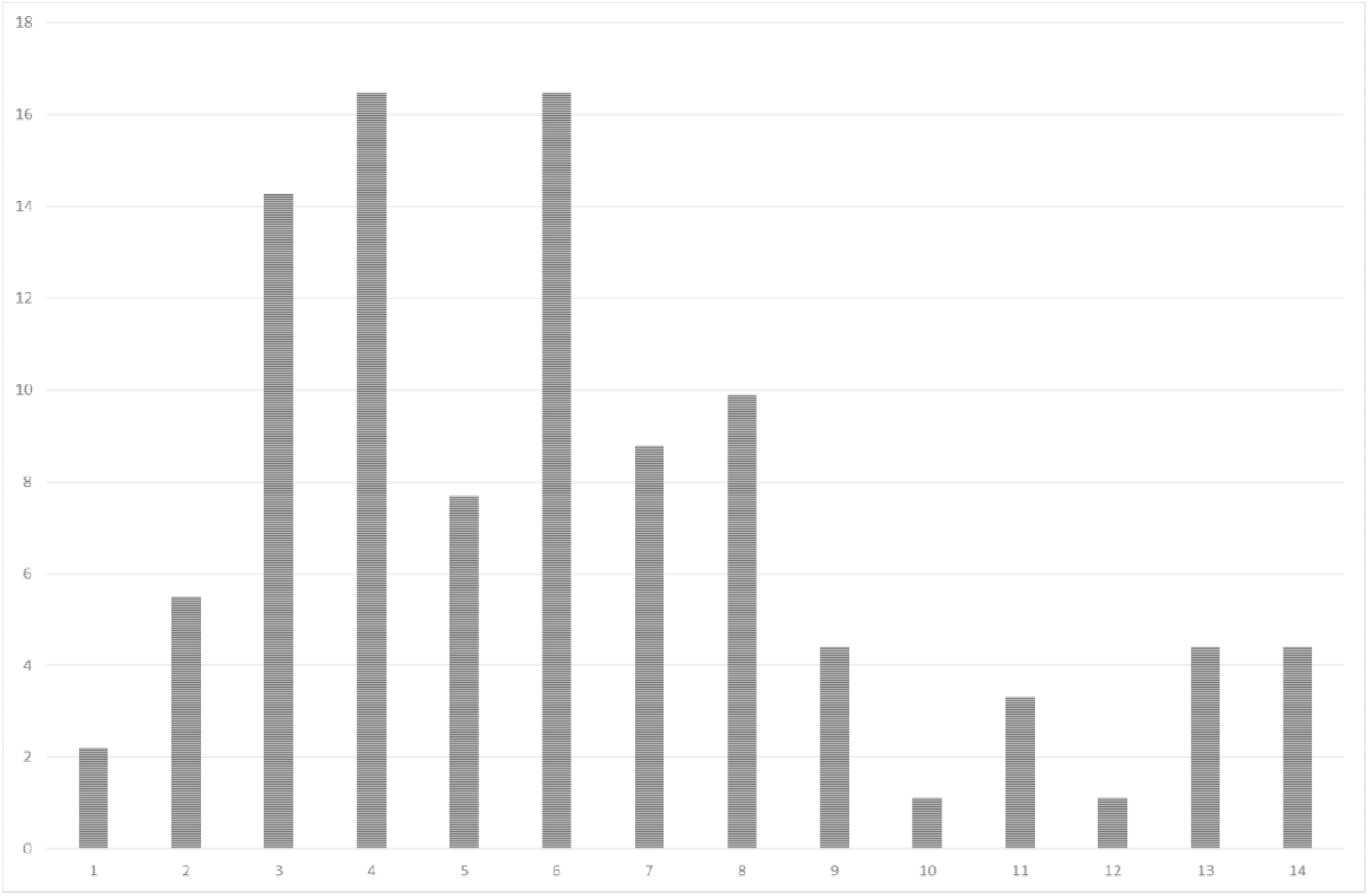
Empirical distribution of the incubation period published by MLHW, Japan. Notes: Bars show the distribution of incubation periods for 91 cases for which the 6 exposure date and onset date were published by MLHW, Japan. The patients for whom 7 incubation was longer than 14 days are included in the bar shown for day 14.

Figure 3 presents data of Apple and Docomo. Their correlation coefficient among was 0.91. It was high, but not extremely high. Estimation results are summarized in the table. The order polynomial function was determined as one except for Docomo data in the whole period as three. The adjusted coefficients of determination, which were used as an index of the goodness of fit, were higher for Docomo data during the entire period. However, after March 10, the adjusted coefficients of determination were better in Apple data.

**Table.**
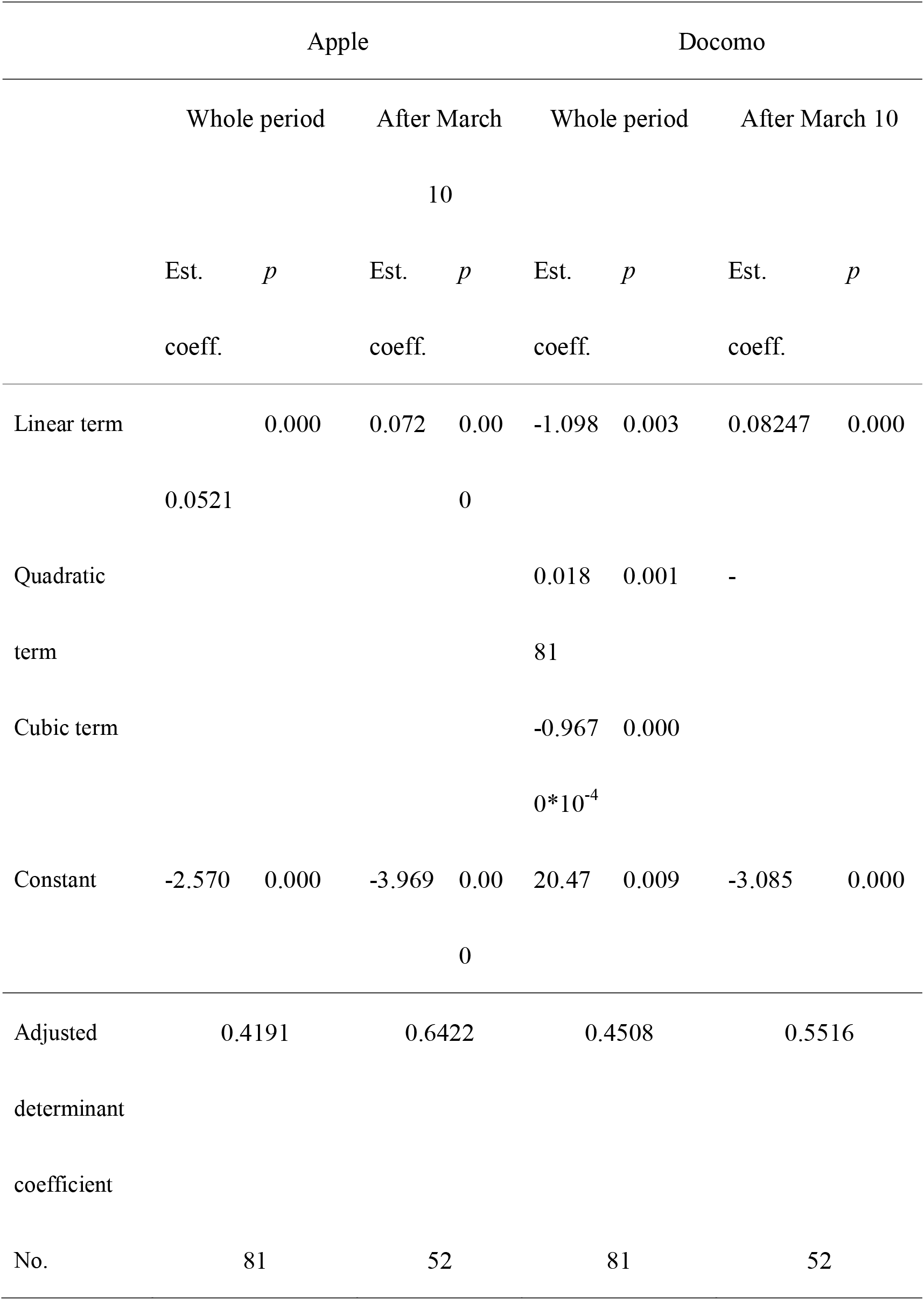
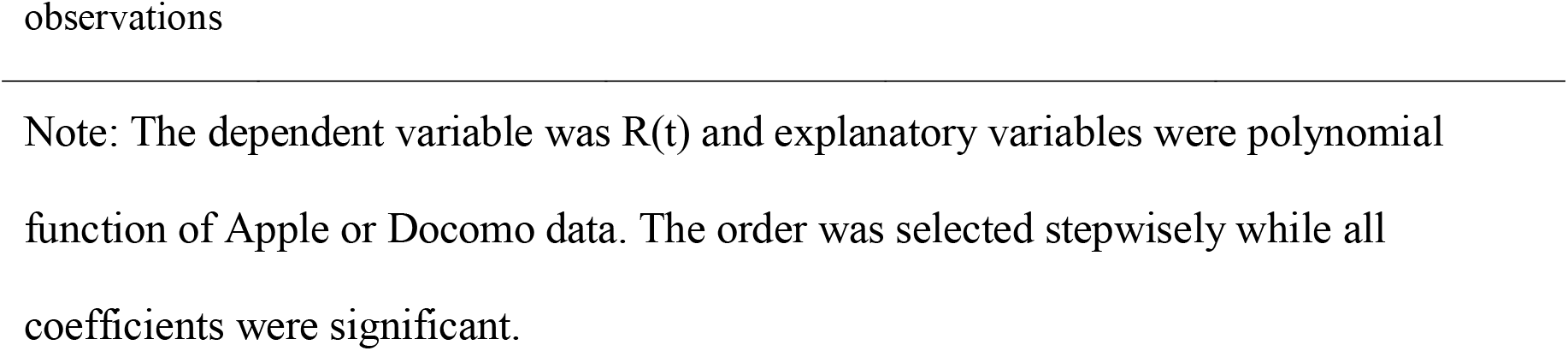
Estimation results of estimate R(t) on the Apple or Docomo data.

|  | Apple |  |  |  | Docomo |  |  |  |
| --- | --- | --- | --- | --- | --- | --- | --- | --- |
|  | Whole period |  | After March |  | Whole period |  | After March 10 |  |
|  |  |  | 10 |  |  |  |  |  |
|  | Est. | *p* | Est. | *p* | Est. | *p* | Est. | *p* |
|  | coeff. |  | coeff. |  | coeff. |  | coeff. |  |
| Linear term |  | 0.000 | 0.072 | 0.00 | -1.098 | 0.003 | 0.08247 | 0.000 |
|  | 0.0521 |  |  | 0 |  |  |  |  |
| Quadratic |  |  |  |  | 0.018 | 0.001 | - |  |
| term |  |  |  |  | 81 |  |  |  |
| Cubic term |  |  |  |  | -0.967 | 0.000 |  |  |
| | | | | | $0*10^{-4}$ | | | |
| Constant | -2.570 | 0.000 | -3.969 | 0.00 | 20.47 | 0.009 | -3.085 | 0.000 |
|  |  |  |  | 0 |  |  |  |  |
| Adjusted | 0.4191 |  | 0.6422 |  | 0.4508 |  | 0.5516 |  |
| determinant |  |  |  |  |  |  |  |  |
| coefficient |  |  |  |  |  |  |  |  |
| No. | 81 |  | 52 |  | 81 |  | 52 |  |

Note: The dependent variable was $R(t)$ and explanatory variables were polynomial function of Apple or Docomo data. The order was selected stepwisely while all coefficients were significant.

**Figure 3:**
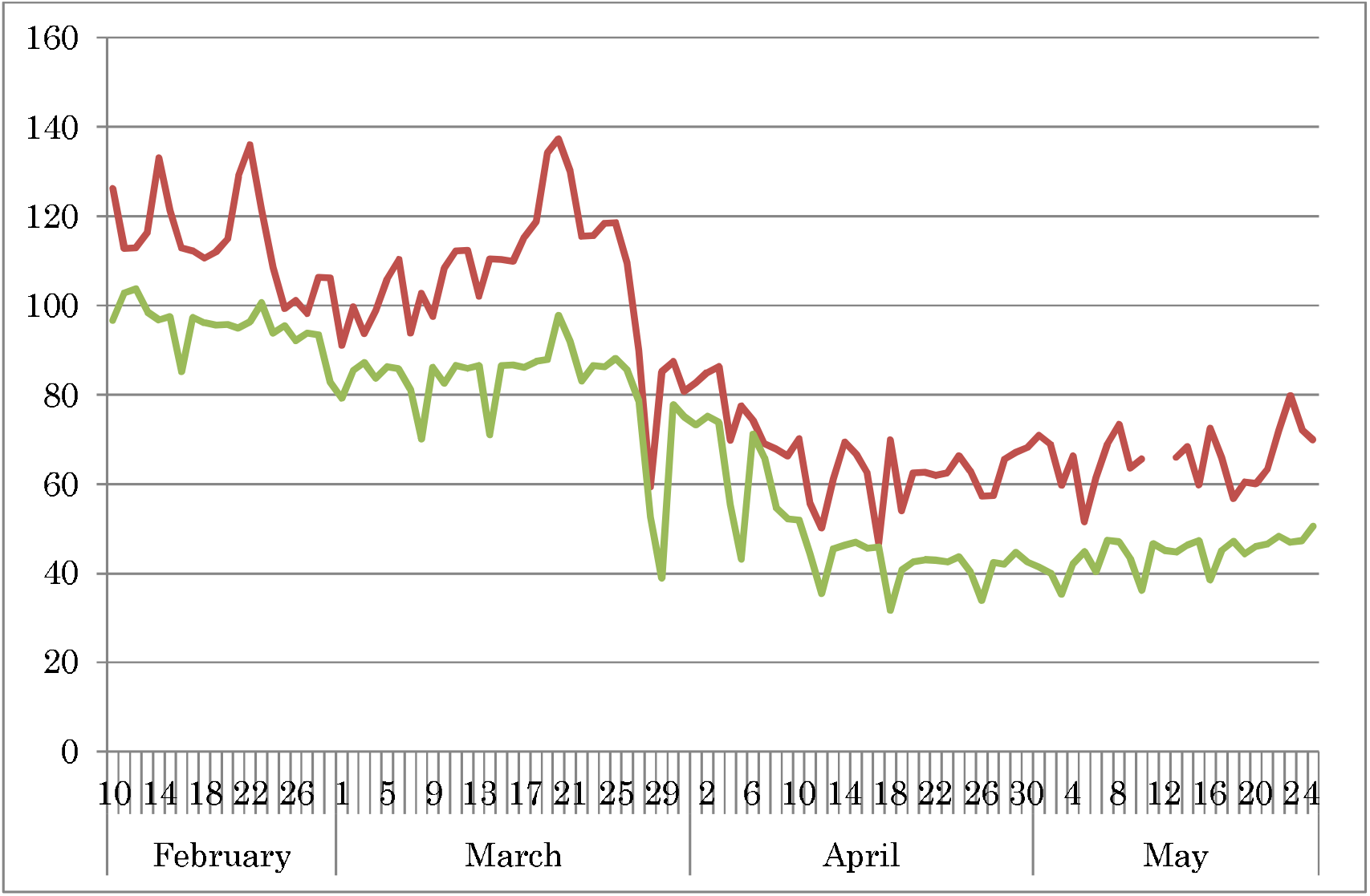
Proportion of going out in Apple and Docomo data. Note: The red line represents the proportion of going out in Apple data comparison with normal level as January 13. The green line represents proportion of going out in Docomo data defined by 500 m mesh.

When we regressed R(t) on a polynomial function of daily Apple and Docomo data simultaneously, the estimated coefficient of Apple data was 0.0648 (*p* = 0.000), but the estimated coefficient of Docomo data was −0.01577 (*p* = 0.386) throughout the whole period. After March 10, for Apple data, it was 0.0640 (*p* = 0.000), but for Docomo data, it was 0.0108 (*p* = 0.627). Therefore, Apple data were significant conditional on Docomo data. However, Docomo data were not significant conditional on Apple data.

Figure 4 depicts the observed R(t) and prediction lines from Apple and Docomo data since March 10. Clearly, neither dataset is able to explain the peak of R(t) around mid-March. Especially, Docomo data were found to have less goodness of fit than Apple data. Conversely, in early April, prediction by Docomo data might overshoot.

**Figure 4:**
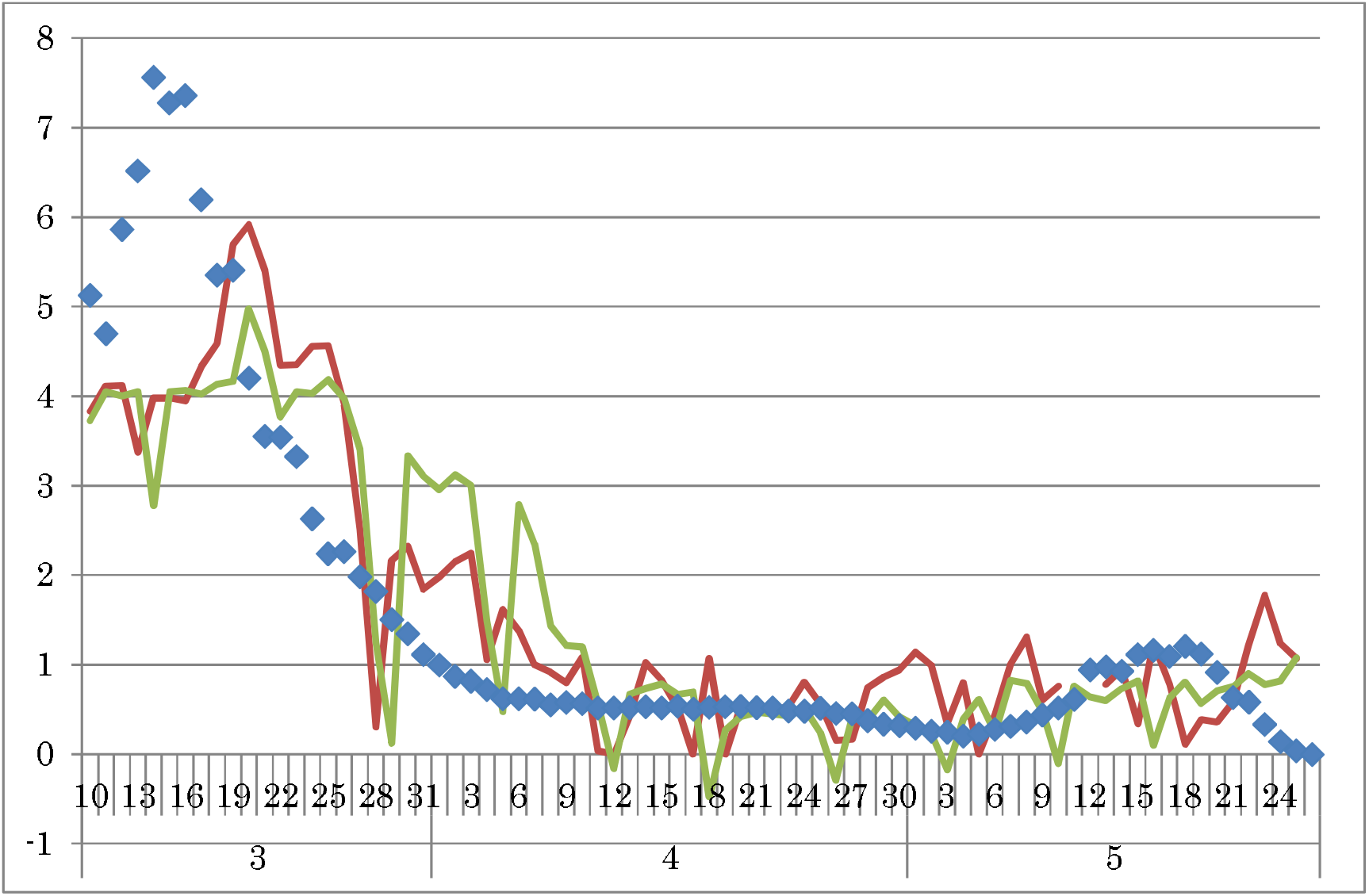
Observed R(t) and the fitted lines from Apple or Docomo data since March 10. Notes: Dots denote the observed R(t). Red line represents the fitted and prediction by Apple data; green line was the line by Docomo data. The data period for retrospective estimation extended to the end of April. The prospective predictions obtained using the two datasets are shown for May.

## Discussion

We showed that Apple data and Docomo data were similar, but Apple data were more informative than Docomo data. Apple data might sufficiently reflect the situation of the COVID-19 outbreak in real time.

The present study has some limitations. First, although we examined the explanatory power for the COVID-19 outbreak nationwide, its applicability at the prefecture level must be verified. Secondly, one must be reminded that Apple and Docomo data indicate the proportion of users in an area who leave their residence. The data do not directly indicate a number of contacts or even a rate of contact. In other words, Apple and Docomo data reflect no intensity of the respective contacts. In fact, such measurement of contact intensity is extremely difficult. Methods to obtain such measurements stand as an objective for future research.

## Data Availability

Japan Ministry of Health, Labour and Welfare. Press Releases. (in Japanese)
Apple. Mobility trend Data (in Japanese)

https://www.mhlw.go.jp/stf/newpage_10723.html

https://www.apple.com/covid19/mobility

## Conclusion

We demonstrated that mobility data from Apple might be better than Docomo data for explaining the entire course of the outbreak in COVID-19 in Japan. Therefore, monitoring Apple data might be sufficient to adjust control measures to maintain the effective reproduction number as less than one. Recently, though it was beyond the study duration of the present study, as of the end of May, Apple data indicate the predicted R(t) as about 1.6. The importance of using Apple data for real time recognition of characteristics of the COVID-19 outbreak is expected to increase.

## Acknowledgments

We acknowledge the great efforts of all staff at public health centers, medical institutions, and other facilities who are fighting the spread and destruction associated with COVID-19.

